# Assessing Depression and Suicidality Among Recently Unemployed Persons with Obstructive Sleep Apnea and Socioeconomic Inequality

**DOI:** 10.1101/2022.04.20.22274101

**Authors:** Vijay Singh, Patricia L. Haynes, Stuart F. Quan

**Affiliations:** Burrell College of Osteopathic Medicine, Las Cruces, NM; Department of Health Promotion Sciences, Mel and Enid Zuckerman College of Public Health, University of Arizona, Tucson, AZ; Division of Sleep and Circadian Disorders, Brigham and Women’s Hospital, Harvard Medical School, Boston, MA; Asthma and Airway Disease Research Center, University of Arizona, Tucson, AZ

**Keywords:** Obstructive Sleep Apnea, Unemployment, Job Loss, Depression, Suicide

## Abstract

**Background:** Obstructive Sleep Apnea (OSA) is a common sleep-related breathing disorder that often is associated with several psychiatric conditions. Job loss is a stressful life event that can also affect mental health and socioeconomic status (SES). We investigated whether there was an association between the prevalence of OSA and several psychiatric conditions within a cohort of persons who recently became unemployed and whether SES was a contributing factor.

**Methods:** Data from 292 participants who completed the screening evaluation of the Assessing Daily Activity Patterns through occupational Transitions (ADAPT) Study were used to assess the association between the prevalence of OSA, and current and past depression, and past suicidality. A type III sleep home sleep monitor was used to identify the presence of OSA and assess its severity. Depression and suicidality were ascertained using the Mini-international neuropsychiatric interview. Years of education was used as a proxy for SES.

**Results:** There were no significant associations between severity of OSA, SES and current depression, past depression, and suicidality. Past suicidality was noted to be more common among those who were single/widowed (17.4%) or those who were divorced or separated (11.1%) (p=0.027). Current depression was more common among Hispanics in comparison to non-Hispanics. Furthermore, prevalence rates of both depression and past suicidality were higher than previous reports in general populations.

**Conclusions:** Within a cohort of individuals who experienced recent job loss, there was no association between OSA and depression or past suicidality. Prevalence rates of both depression and past suicidality were higher than previous reports in the general population.

## Introduction

Obstructive sleep apnea (OSA) is a highly prevalent medical condition characterized by recurrent episodes of partial or complete obstruction of the upper airways during sleep. Common symptoms are loud snoring, witnessed apneas, and daytime fatigue or sleepiness (Young et al., 1993). It is associated with various mental health conditions such as depression, anxiety, and post-traumatic stress disorder. Importantly, recent studies with small sample sizes have shown a correlation between suicidal behavior and persons with OSA (Choi et al., 2015).

Additionally, OSA has also been shown to relate to lower socioeconomic status (SES) which can be impacted by job loss (Guglielmi et al., 2019). However, the association of OSA, SES with mental health conditions in a population of individuals who have been recently unemployed has not adequately explored.

In this study, we evaluated the association between OSA and SES and a self-reported history of several common psychiatric disorders. We hypothesized that individuals with recent job loss and OSA would have an increased likelihood of having a history of self-reported psychiatric illness and that SES would be contributing factor.

## Methods

Study participants were part of the ADAPT Study, an 18-month longitudinal study examining changes in sleep, social rhythms, and obesity following an involuntary job loss. The study protocol and recruitment strategy have been described in detail previously (Haynes et al., 2017). Briefly, all individuals who applied for unemployment insurance (UI) in the greater Tucson, Arizona and surrounding areas between October 2015 and December 2018 received study recruitment flyers within their UI intake packets. Interested individuals contacted study staff and completed phone screens assessing exclusion criteria; potentially eligible individuals were then scheduled for in-person screening visits. Individuals were eligible for overnight screening if they had experienced an involuntary job loss within 90 days of study enrollment, had been with their employer for at least six months, were currently employed less than five hours per week and did not complete any night shift work within the last 30 days. During the in-person screening, participants provided written informed consent, as well as information about their demographics, employment, and medical history. They also were screened for homelessness, existing physiological and mental health conditions, substance abuse, and major sleep diagnoses which could interfere with social rhythms and sleep patterns. An overnight at-home screening for sleep apnea was performed utilizing the ApneaLink Plus™ (ResMed, San Diego, CA) to exclude moderate sleep apnea as a cause of sleep disruption. This study was approved by the University of Arizona Human Subjects Protection Program (#1401185772).

Data used in this analysis originated from the study’s screening visit. Of the 446 adults who provided written consent, 292 adult participants underwent overnight screening for obstructive sleep apnea (OSA) and completed the Mini-international neuropsychiatric interview (Sheehan et al., 1998). Participants were classified as having a current or past diagnosis of several mental health disorders including depression, mania, hypomania and suicidality on the basis of their responses to the structured screening interview. The screening survey also recorded information pertaining to socioeconomic status, educational attainment, race, and ethnicity. Severity of OSA was based on the Apnea-Hypopnea Index (AHI) from the home sleep study as follows: No OSA (AHI <5 /h), Mild OSA (AHI ≥5 and <15 /h), Moderate to Severe OSA (AHI ≥15 /h). Primary dichotomous endpoints included percent of people who had past or current episodes of depression, and past episodes of suicidality. Income in relation to the US Health and Human Services Poverty Line for Arizona was used as an indicator of SES based on data from the baseline survey.

### Statistical Analysis

Mean and standard deviation for the variables being investigated were used to describe the baseline characteristics of the cohort. Comparison of the prevalence of various mental health conditions as a function of the presence of OSA was performed using a Pearson-Chi Square test. Comparisons of the prevalence of depression and suicidality to previously reported prevalence rates were completed using the binomial test. All statistical analyses were done using the IBM SPSS version 28 (Armonk, NY).

## Results

Table 1 describes the overall demographic and baseline characteristics for the participants who volunteered to be a part of the survey. Most participants were White and 35.3% were Hispanic. The majority were women and were not married or had a live-in partner. A majority were living at or below the poverty line and two-thirds had a high school education or less. On average, the population being studied was obese. Moderate to severe OSA was observed in 48 participants (16.4%). Screening for mental health conditions showed that approximately one-third of the cohort had self-reported current as well as past depression. Both current and past depression rates are significantly higher (binomial test, p<0.001) than the 8.1% prevalence of depression reported from the 2013-2016 National Health and Nutrition Examination Survey (NHANES) (Brody et al., 2018). History of suicidality was present in approximately 11% of the sample. This rate is higher than the 4.2% prevalence of suicidal thoughts in Arizona reported by the National Survey on Drug Use and Health in 2008-2009 (Crosby et al., 2011)(binomial test, p<0.001). The remaining health conditions were observed in very small numbers and were not analyzed further.

**Table 1:** Baseline Characteristics.

| VARIABLES | N | Mean/% | sd |
| --- | --- | --- | --- |
| *Demographic/Anthropometric* |  |  |  |
| Age (years) | 292 | 41.2 | 10.3 |
| Gender (% Men) | 126 | 43.2 | -- |
| Race |  |  |  |
| % White | 198 | 67.8 | -- |
| % Other Race | 94 | 32.2 | -- |
| Ethnicity |  |  |  |
| % Not Hispanic/Latino | 189 | 64.7 | -- |
| % Hispanic | 103 | 35.3 | -- |
| BMI (kg/m<sup>2</sup>) | 213 | 30.5 | 7.9 |
| Marital Status (% Married/Live-in Partner) | 127 | 43.5 | -- |
| Socioeconomic Status |  |  |  |
| % Less than High School or High School | 193 | 66.1 | -- |
| % Income ≤ 100% of Poverty Line | 164 | 56.2 | -- |
| *Obstructive Sleep Apnea* |  |  |  |
| AHI (No OSA <5) | 163 | 55.8 | -- |
| AHI (Mild 5-15) | 81 | 27.7 | -- |
| AHI (Moderate to Severe >15) | 48 | 16.4 | -- |
| *Mental Health Conditions* |  |  |  |
| Depression (Current) | 95 | 32.5 | -- |
| Depression (Past) | 112 | 38.4 | -- |
| Suicidality (Past) | 33 | 11.3 | -- |
| Mania | 1 | 0.3 | -- |
| Hypomania | 1 | 0.3 | -- |
| Panic Disorder | 22 | 7.5 | -- |
| Social Anxiety Disorder | 3 | 1.0 | -- |
| Agoraphobia | 3 | 1.0 | -- |
| Obsessive Compulsive Disorder | 8 | 2.7 | -- |

Table 2 shows relationships between severity of AHI and marital status, educational attainment, race, and socioeconomic status. No significant associations were observed.

**Table 2:**
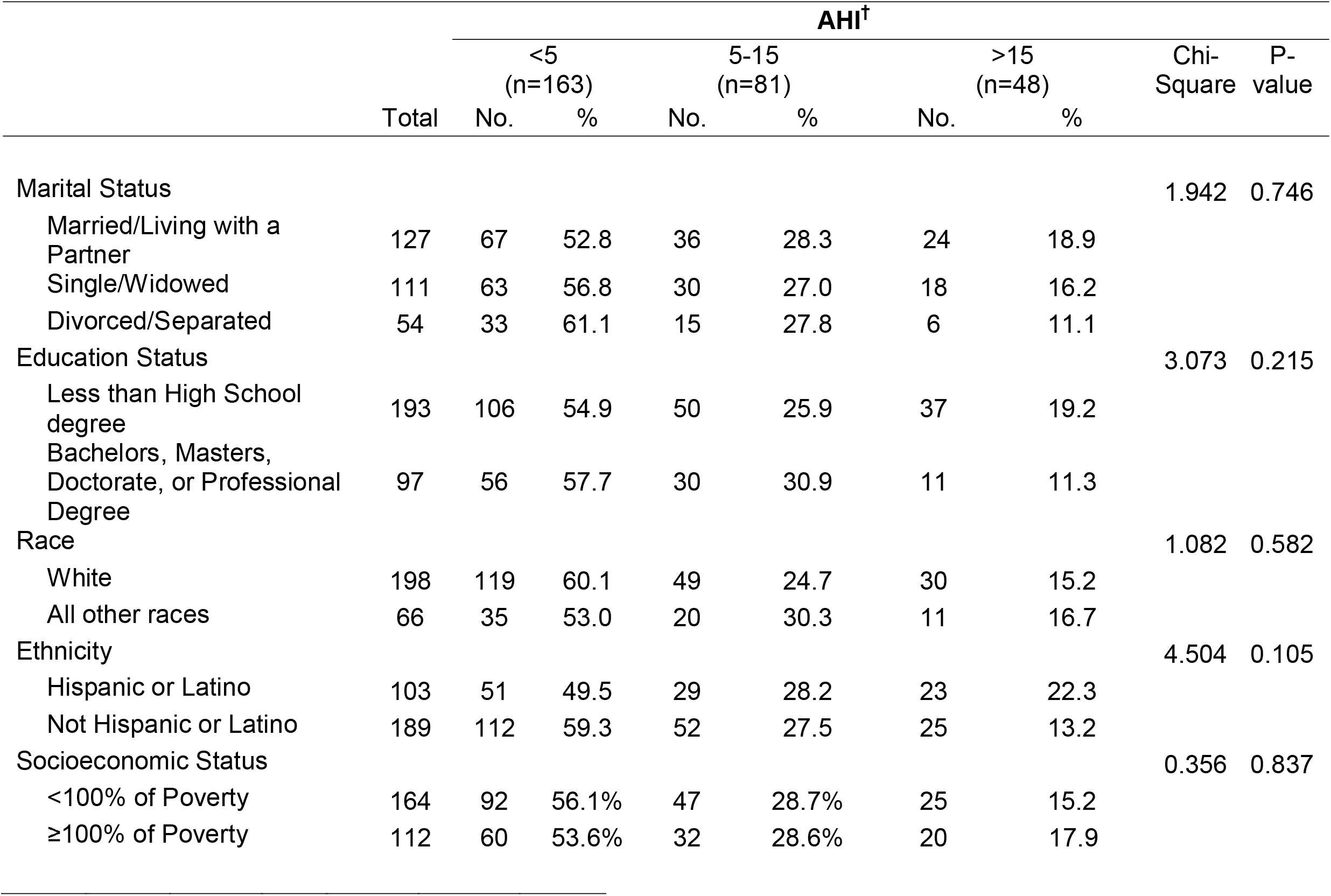
Demographic and Health Condition Characteristics of the Sample by OSA Severity.

Shown in Table 3 are the associations of severity of OSA, social and demographic characteristics of the participants with current and past episodes of depression. There were no significant associations with respect to self-reported current or past depression with any demographic characteristics or severity of OSA.

**Table 3:** Associations of Demographic and Health Conditions With Self-Reported Current and Past Depression.

|  | Current Depression |  |  |  |  |  | Past Depression |  |  |  |  |  |  |  |
| --- | --- | --- | --- | --- | --- | --- | --- | --- | --- | --- | --- | --- | --- | --- |
|  | Total | No Current Depression (n=197) |  | Current Depression (n=95) |  | Chi-Square | P-Value | Total | No Past Depression (n=180) |  | Past Depression (n=112) |  | Chi-Square | P-Value |
|  |  | No. | % | No. | % |  |  |  | No. | % | No. | % |  |  |
| Gender |  |  |  |  |  | 0.570 | 0.450 |  |  |  |  |  | 3.171 | 0.075 |
| Male | 126 | 88 | 69.8 | 38 | 30.2 |  |  | 126 | 85 | 67.5 | 41 | 32.5 |  |  |
| Female | 166 | 109 | 65.7 | 57 | 34.3 |  |  | 166 | 95 | 57.2 | 71 | 42.8 |  |  |
| Age |  |  |  |  |  | 5.128 | 0.274 |  |  |  |  |  | 5.128 | 0.274 |
| 20-29 | 42 | 21 | 11.7 | 21 | 18.8% |  |  | 42 | 21 | 50.0 | 21 | 50.0 |  |  |
| 30-39 | 100 | 61 | 33.9 | 39 | 34.8 |  |  | 100 | 61 | 61.0 | 39 | 39.0 |  |  |
| 40-49 | 75 | 45 | 25.0 | 30 | 26.8 |  |  | 75 | 45 | 60.0 | 30 | 40.0 |  |  |
| 50-59 | 71 | 50 | 27.8 | 21 | 18.8% |  |  | 71 | 50 | 70.4 | 21 | 29.6 |  |  |
| 60+ | 4 | 3 | 1.7 | 1 | 0.9 |  |  | 4 | 3 | 75.0 | 1 | 25.0 |  |  |
| Marital Status |  |  |  |  |  | 1.695 | 0.429 |  |  |  |  |  | 0.521 | 0.771 |
| Married/Living with a Partner | 127 | 90 | 70.9 | 37 | 29.1 |  |  | 127 | 79 | 62.2 | 48 | 37.8 |  |  |
| Single/Widowed | 111 | 74 | 66.7 | 37 | 33.3 |  |  | 111 | 70 | 63.1 | 41 | 36.9 |  |  |
| Divorced/Separated | 54 | 33 | 61.1 | 21 | 38.9 |  |  | 54 | 31 | 57.4 | 23 | 42.6 |  |  |
| Education Status |  |  |  |  |  | 2.346 | 0.126 |  |  |  |  |  | 0.014 | 0.906 |
| Less than High School degree | 193 | 124 | 64.2 | 69 | 35.8 |  |  | 193 | 118 | 61.1 | 75 | 38.9 |  |  |
| Bachelors, Masters, Doctorate, or Professional Degree | 97 | 71 | 73.2 | 26 | 26.8 |  |  | 97 | 60 | 61.9 | 37 | 38.1 |  |  |
| Race |  |  |  |  |  | 0.212 | 0.645 |  |  |  |  |  | 1.545 | 0.214 |
| White | 198 | 135 | 68.2 | 63 | 31.8 |  |  | 198 | 118 | 59.6 | 80 | 40.4 |  |  |
| Underrepresented Minority | 66 | 47 | 71.2 | 19 | 28.8 |  |  | 66 | 45 | 68.2 | 21 | 31.8 |  |  |
| Ethnicity |  |  |  |  |  | 7.519 | 0.006 |  |  |  |  |  | 1.281 | 0.258 |
| Hispanic or Latino | 103 | 59 | 57.3 | 44 | 42.7 |  |  | 103 | 59 | 57.3 | 44 | 42.7 |  |  |
| Not Hispanic or Latino | 189 | 138 | 73.0 | 51 | 27.0 |  |  | 189 | 121 | 64.0 | 68 | 36.0 |  |  |
| Socioeconomic Status |  |  |  |  |  | 1.923 | 0.165 |  |  |  |  |  | 0.000 | 0.997 |
| <100% of Poverty | 164 | 104 | 63.4 | 60 | 36.6 |  |  | 164 | 101 | 61.6 | 63 | 38.4 |  |  |
| ≥100% of Poverty | 112 | 80 | 71.4 | 32 | 28.6 |  |  | 112 | 69 | 61.6 | 43 | 38.4 |  |  |
| Apnea Hypopnea Index |  |  |  |  |  | 1.641 | 0.441 |  |  |  |  |  | 3.359 | 0.186 |
| Less than 5 | 163 | 115 | 70.6 | 48 | 29.4 |  |  | 163 | 95 | 58.3 | 68 | 41.7 |  |  |
| ≥5 and <15 | 81 | 52 | 64.2 | 29 | 35.8 |  |  | 81 | 50 | 61.7 | 31 | 38.3 |  |  |
| ≥15 | 48 | 30 | 62.5 | 18 | 37.5 |  |  | 48 | 35 | 72.9 | 13 | 27.1 |  |  |

The associations between severity of OSA and demographic characteristics with a history of self-reported past suicidality are shown in Table 4. Past suicidality was least likely to have occurred among those who were married or had a live-in partner (6.6%) in comparison to those who were single/widowed (17.4%) or those who were divorced or separated (11.1%) (p=0.027). No other significant relationships were found between these demographic factors as well as history of OSA.

**Table 4:** Associations of Demographic and Health Conditions With Past History of Suicidality.

|  | Past Suicidality |  |  |  |  | Chi-Square | P-Value |
| --- | --- | --- | --- | --- | --- | --- | --- |
|  | Total | No Past Suicidality (n=257) |  | Past Suicidality (n=33) |  |  |  |
|  |  | No. | % | No. | % |  |  |
| Gender |  |  |  |  |  | 0.249 | 0.618 |
| Male | 126 | 113 | 89.7% | 13 | 10.3% |  |  |
| Female | 164 | 144 | 87.8% | 20 | 12.2% |  |  |
| Age |  |  |  |  |  | 2.966 | 0.563 |
| 20-29 | 41 | 34 | 81.0% | 7 | 16.7% |  |  |
| 30-39 | 100 | 87 | 87.0% | 13 | 13.0% |  |  |
| 40-49 | 74 | 67 | 89.3% | 7 | 9.3% |  |  |
| 50-59 | 71 | 65 | 91.5% | 6 | 8.5% |  |  |
| 60+ | 4 | 4 | 100.0% | 0 | 0.0% |  |  |
| Marital Status |  |  |  |  |  | 7.213 | 0.027 |
| Married/Living with a Partner | 127 | 119 | 93.7% | 8 | 6.6% |  |  |
| Single/Widowed | 109 | 90 | 82.6% | 19 | 17.4% |  |  |
| Divorced/Separated | 54 | 48 | 88.9% | 6 | 11.1% |  |  |
| Education Status |  |  |  |  |  | 0.616 | 0.433 |
| Less than High School degree | 192 | 172 | 89.6% | 20 | 10.4% |  |  |
| Bachelors, Masters, Doctorate, or Professional Degree | 96 | 83 | 86.5% | 13 | 13.5% |  |  |
| Race |  |  |  |  |  | 0.635 | 0.425 |
| White | 196 | 171 | 87.2% | 25 | 12.8% |  |  |
| All other races | 66 | 60 | 90.9% | 6 | 9.1% |  |  |
| Ethnicity |  |  |  |  |  | 0.175 | 0.244 |
| Hispanic or Latino | 101 | 93 | 92.1 | 8 | 7.9 |  |  |
| Not Hispanic or Latino | 189 | 164 | 86.8 | 25 | 13.2 |  |  |
| Socioeconomic Status |  |  |  |  |  | 1.649 | 0.199 |
| <100% of Poverty | 162 | 141 | 87.0% | 21 | 13.0% |  |  |
| ≥100% of Poverty | 112 | 103 | 92.0% | 9 | 8.0% |  |  |
| Apnea Hypopnea Index |  |  |  |  |  | 0.107 | 0.948 |
| Less than 5 | 162 | 144 | 88.9% | 18 | 11.1% |  |  |
| ≥5 and <15 | 81 | 72 | 88.9% | 9 | 11.1% |  |  |
| ≥15 | 47 | 41 | 87.2% | 6 | 12.8% |  |  |

## Discussion

In this study of recently unemployed adults, we did not find any associations between OSA or SES with the prevalence of self-reported depression (past or current) or past suicidality. However, the prevalence rates for depression were higher in this cohort in comparison to previous reports in the general population particularly among Hispanics. History of suicidality was also higher than reported in the general Arizona population and was observed to be more common among those who were not married or living with a partner.

Although we were unable to find any associations between OSA and psychiatric conditions in our study, most but not all previous literature has found correlations between OSA and patients suffering from depression (Ejaz et al., 2011; Hobzova et al., 2017). There are many similarities between OSA and depression including symptoms such as poor sleep, anhedonia, low energy, and lack of concentration (Jehan et al., 2017). It is unclear why our results do not confirm previous findings of a close relationship. However, our cohort had relatively few participants with severe OSA, as compared to samples from sleep centers. It is possible that depressive symptoms are correlated with the AHI only in severe OSA (Lee 2015). Additionally, although most individuals in the study were female who are more likely to have major depression (Albert, 2015), the prevalence of OSA is higher in males (Vanek et al., 2020). Thus, the ability to detect an association between depression and OSA may be low. Furthermore, the prevalence of current (30.2% Male/34.3% Female) and past depression (32.5% Male/42.8% Female) was already high in our cohort of recently unemployed persons in comparison to estimates of the overall prevalence before the COVID-19 pandemic of ∼8% (Brody et al., 2018; Cao et al., 2020). Thus, it is possible that any additional impact of OSA would be difficult to detect.

Increased rates of depression have been previously reported among unemployed adults (Gallup Inc, 2014). We also found an elevated rate of depression among unemployed persons, but extend these findings to specifically those with recent job loss. Although our depression prevalence rate was higher than previously observed (Gallup Inc, 2014), this comparison should be interpreted cautiously given the dissimilarities in ascertainment instruments. It might be surmised that recent job loss is the most plausible explanation for high rates of current depression in this cohort, but it is unclear why the prevalence of past depression also is high. One explanation is that history of depression is a marker for subsequent unemployment as has been previously suggested (Andreeva et al., 2015). Irrespective of etiology, our results indicate that recently unemployed persons are at high risk for mental health issues.

Hispanics in our study had higher rates of current depression than non-Hispanics. In a review of depression in Hispanics in the United States, Lewis-Fernandez et al (2005) noted that initial studies found that Hispanic populations have higher rates of depression than primarily white populations, but that this was not confirmed in later studies. Hispanics are a heterogenous ethnic group and are primarily of Mexican ancestry in Arizona. In their review, Lewis-Fernandez et al (2005) noted that United States-born Mexican Americans had higher rates of depression than those born in Mexico. Although ability to speak English was an inclusion requirement for participation, we could not precisely determine the geographic origin of the Hispanics in this cohort. Thus, the explanation for our finding of higher depression rates in Hispanics remains unclear.

Our study did not find an association between OSA and past suicidality. In contrast, a recent large Danish cohort study of 48,168 patients with OSA found an increased risk of suicide as well as self-harm (Udholm et al., 2022). It is likely that we were underpowered to detect any relationship, since only 33 participants reported past suicide. However, we did find a history of suicidality to be greater in those were single or widowed. This is consistent with previous studies demonstrating suicide rates are higher amongst those who are not married (Mastekaasa, 1995). Our data provide additional evidence showing that suicidality is more common among individuals who do not cohabitate.

Similar to our findings related to the prevalence of depression, the past suicidality rate in our cohort was higher than reported in general populations (Choi et al., 2015; Crosby et al., 2011; Kessler et al., 2005). This is most likely related to higher rates of past depression in our cohort.

Lower SES has been associated with a higher occurrence of psychiatric co-morbidities. Patients with lower SES have been noted to have a greater likelihood of developing chronic mental health conditions, particularly depression (Lorant et al., 2003). Studies of OSA and health disparities have demonstrated a higher prevalence of OSA among those who are disadvantaged (Etindele Sosso & Matos, 2021). Although a majority of the cohort had incomes below the poverty line, this status may have been a recent event associated with job loss and therefore not sufficiently impactful to increase the prevalence of OSA.

Our study has several strengths. We assessed a moderate sized cohort consisting of 292 individuals, 126 of which were of the male gender. This allowed us to have a nearly balanced analysis of both male and female genders. Moreover, it is unique in that we explored associations of OSA and mental health among those who were recently unemployed. However, there are some limitations. In particular, our cohort does not reflect the ethnic distribution of the general US population. Approximately one-third of the cohort was Hispanic which is approximately twice the proportion in the US, but nevertheless reflects the ethnic composition of Tucson, AZ. In addition, the relative paucity of persons with severe OSA may have limited our power to detect significant associations.

In conclusion, in this study, we did not find any notable relationships between OSA, SES and depression and past suicidality in those who were recently unemployed although rates of both conditions were higher than previously reported in the general population. Further studies will need to be undertaken with a larger more diverse population to ascertain to confirm these findings.

## Data Availability

All data produced in the present study are available upon reasonable request to the authors

## Acknowledgements

This research was supported by the National Heart, Lung, and Blood Institute (#1R01HL117995-01A1; PI: Haynes).

